# Exploring the association between food insecurity and overweight/obesity among adults in low- and middle-income countries: A systematic review protocol

**DOI:** 10.1101/2025.07.30.25332450

**Authors:** Jonathan Lara-Arevalo, Layla White, Alyssa Walker-Porter, Shu Wen Ng, Lamis Jomaa, Lindsey Smith Taillie

## Abstract

**Background:** In low- and middle-income countries (LMICs), food insecurity has traditionally been associated with undernutrition. However, as many LMICs undergo a rapid nutrition transition, greater access to inexpensive, energy-dense, and nutrient-poor foods is contributing to rising rates of overweight and obesity. These shifts may be altering the relationship between food insecurity and excess weight, a link that has been more widely documented in high-income countries. This systematic review aims to synthesize evidence on the association between food insecurity and overweight/obesity among adults in LMICs, and to identify proposed mechanisms and contextual factors that may influence this relationship.

**Methods:** Primary observational studies focused on the association between food insecurity and overweight/obesity among adults in LMICs will be systematically searched from multiple electronic databases, including PubMed, EMBASE, Global Health, Scopus, and LILACS. Grey literature will be explored using Google Scholar and ProQuest Dissertations & Theses Global. Reference lists of included studies and relevant reviews will also be manually screened to identify additional studies. Extracted data will include measures of food insecurity, overweight/obesity outcomes, and any reported mechanisms or contextual factors influencing their relationship.

**Discussion:** This review will contribute to a deeper understanding of the complex and evolving relationship between food insecurity and overweight/obesity in LMICs. Where data allow, we will explore how this association varies across country income levels, urban and rural settings, sex, age groups, and exposure to extreme events. Identified mechanisms and contextual modifiers will be synthesized to provide insights into the pathways that may underlie these associations. The findings will help identify knowledge gaps and inform future research and policy efforts aimed at addressing the dual burden of malnutrition in LMICs.

**Registration:** This systematic review protocol is registered in PROSPERO under the registration number: CRD420251076653.

## Introduction

Food insecurity—defined as the condition in which people lack reliable access to sufficient, safe, and nutritious foods for normal growth, development, and a healthy life—is a major global concern, particularly in low- and middle-income countries (LMICs) (1, 2). In 2023, nearly 30% of the world population was moderately or severely food insecure, with 864 million individuals experiencing severe levels, the majority living in LMICs (2). Food insecurity in these settings is primarily driven by limited household purchasing power and is exacerbated by climate change, poverty, economic inequities, and armed conflict, which disrupt food systems and reduce availability and access to food (3-10).

Traditionally, food insecurity in LMICs has been linked to hunger and undernutrition. Adults experiencing food insecurity in these settings often have lower diet quality, with limited access to nutrient-dense foods (11, 12). Studies have consistently documented associations between food insecurity and indicators of undernutrition, such as stunting, wasting, and underweight (11, 13-15). These outcomes are influenced by structural factors including limited financial resources, poor proximity to nutritious foods, and lack of access to safe water, health services, transportation or refrigeration (11, 13).

However, this paradigm has been shifting with many LMICs undergoing remarkable nutrition transition characterized by increased consumption of diets high in refined foods and nutrients of concern, such as saturated fats, sugars, and sodium (16-18). As a result, these countries are now facing what is commonly referred to as the triple burden of malnutrition, in which undernutrition, overnutrition, and micronutrient deficiencies coexist within the same population (19). This global nutrition transition has made energy-dense, nutrient-poor foods more affordable and accessible, especially among low-income households (17, 20). Aggressive marketing strategies by the food and beverage industry targeting low-income populations have also contributed to the increased consumption of highly processed, energy-dense food and beverage products (21) that frequently contain many additives to make them highly palatable and some argue addictive (22). Consequently, food-insecure individuals may increasingly rely on highly processed foods and diets rich in nutrients of concern and low in nutrients of benefit, potentially heightening their risk of overweight and obesity (23, 24).

In contrast to LMICs, the relationship between food insecurity and overweight/obesity has been extensively documented in high-income countries (HICs), where it is often referred to as the “food insecurity-obesity paradox” (25-27). In these settings, the availability of cheap, ultra-processed foods, combined with limited access to healthier options in socioeconomically disadvantaged neighborhoods, has contributed to rising obesity rates, particularly among food insecure populations (25, 28). This interconnection between constrained access and unhealthy food environments highlights the complex pathways through which food insecurity may lead to excess weight.

Although LMICs are increasingly vulnerable to similar dynamics, evidence examining the association between food insecurity and overweight/obesity in these contexts remains limited. This is particularly concerning given the limited healthcare infrastructure and resources necessary to prevent and manage the growing burden of diet and obesity-related diseases in LMICs (23). Existing reviews on food insecurity and nutritional outcomes have primarily focused on undernutrition, or have assessed multiple weight-related outcomes without a specific focus on overweight/obesity in LMICs contexts (29, 30). Although former reviews provided important foundational insights, they were published over seven years ago and therefore do not encompass the substantial and rapidly expanding body of research on this topic. (23, 29, 30).

To address these gaps, this systematic review will evaluate the association between food insecurity and overweight/obesity among adults (aged 18 years and older) living in LMICs. It will also describe proposed mechanisms, contextual factors, and research gaps, contributing to a greater understanding of this emerging relationship and informing future research and policy efforts in LMICs.

## Methods

This protocol was developed in accordance with the Preferred Reporting Items for Systematic Reviews and Meta-Analyses (PRISMA) 2020 guidelines (31). A completed PRISMA for Protocols (PRISMA-P) checklist (32) is provided in the **S1 file**.

### Eligibility Criteria

This review will include empirical observational studies that assess food insecurity and include overweight/obesity as outcome measures. Eligible designs include cross-sectional, cohort, and case-control studies. Observational studies are appropriate for this review because food insecurity is a real-world exposure shaped by complex socioeconomic and structural factors and cannot be ethically randomized. While some randomized controlled trials (RCTs) may evaluate interventions aiming to reduce food insecurity, they are not designed to assess the association between food insecurity and overweight/obesity. For this reason, RCTs will be excluded, along with review articles, meta-analyses, expert opinions, commentaries, letters to the editor, and articles without quantitative data or insufficient methodological detail.

The review will be limited to studies published from 2000 onward, as the global context has shifted substantially over the past 25 years and more recent evidence is considered more relevant to current policy and epidemiological trends. Eligible studies must be conducted in LMICs, as defined by the 2024-2025 World Bank’s income classification based on gross national income per capita (33). This includes countries classified as low-income, lower-middle-income, and upper-middle-income. Eligible studies must report data collected from populations residing in LMICs during the study period. Multinational studies will be included only if data for LMICs are reported separately or can be extracted independently.

Included studies must focus on adults aged 18 years or older. In studies that include both adult and child populations, only the data pertaining to adults will be analyzed. Studies will be excluded if they assess food insecurity without any accompanying measure of body composition or anthropometric indicators, or if they focus exclusively on populations with medical conditions that substantially alter body composition (e.g., advanced stages of infectious diseases such as HIV/AIDS). Studies with fewer than 30 participants also be excluded.

To be eligible, studies must evaluate food insecurity at the household or individual level using validated experience-based food security scales. Examples include the Household Food Insecurity Access Scale (HFIAS) (34), Latin American and Caribbean Food Security Scale (ELCSA) (35), Food Insecurity Experience Scale (FIES) (36), Brazilian Food Insecurity Scale (EBIA) (37), and other instruments adapted from the United States Department of Agriculture Food Security Survey Module (USDA-FSSM) (38), among others. These experiential scales capture lived experiences and behaviors related to constrained food access due to limited resources (39). A summary of the inclusion and exclusion criteria is provided in **Table 1**.

**Table 1.** Inclusion and exclusion criteria of studies.

| Characteristics | Inclusion Criteria | Exclusion Criteria |
| --- | --- | --- |
| Study Characteristics | Observational studies, including cross-sectional, cohort, and case-control studies | Randomized controlled trials (RCTs), review articles, meta-analyses, expert opinions, commentaries, letters to the editor, and articles without quantitative data or insufficient methodological detail |
|  | Published from the year 2000 onward | Published before the year 2000 |
|  | Studies conducted in low- and/or middle-income countries | Studies conducted in high-income countries |
|  | Written in English or Spanish language | Written in other languages than English or Spanish |
| Population | Studies that assess adult individuals (aged 18 years or older) | Studies that only assess children aged less than 18 years |
|  | Studies with more than 30 participants | Studies with fewer than 30 participants |
|  |  | Studies conducted exclusively among individuals with morbid conditions that severely affect body composition (e.g., advanced infectious diseases such as HIV/AIDS) |
| Exposure | Studies using a validated | Studies not using a validated |
|  | instrument to measure food insecurity | instrument to measure food insecurity |
|  |  | Studies assessing only a single component of food insecurity (e.g., food access) |
| Outcome | Studies that assess both food insecurity and body composition (e.g., overweight, obesity, BMI, waist circumference) | Studies that assess only food insecurity without any measure of body composition or anthropometric indicators |

### Information sources and search strategy

The following databases will be searched: PubMed, EMBASE, Global Health, Scopus, and LILACS. To capture relevant grey literature, additional searches will be conducted using Google Scholar and ProQuest Dissertations & Theses Global. To ensure comprehensiveness, the reference lists of all included studies and relevant reviews (including narrative, scoping, and/or systematic reviews) will be manually screened to identify additional eligible studies not captured through database searches.

The search strategy was developed for each database with the assistance of a research librarian at the University of North Carolina at Chapel Hill. Keywords and controlled vocabulary terms related to “food insecurity”, “overweight”, “obesity”, and “low- and middle-income countries” were used. These terms were identified through preliminary scoping searches, review of relevant systematic reviews, and consultation with the librarian to ensure comprehensive and database-specific coverage. Although the review focuses on adult populations, the term “adult” was excluded from the search strategy to avoid inadvertently omitting studies conducted in adults but not explicitly labeled as such. The search terms used in the search are presented in **Table 2** and the search strategy for all databases is presented in **S2 File** (English) and **S3 File** (Spanish). Searches will be limited to studies published from 2000 onward, in English or Spanish.

**Table 2.**
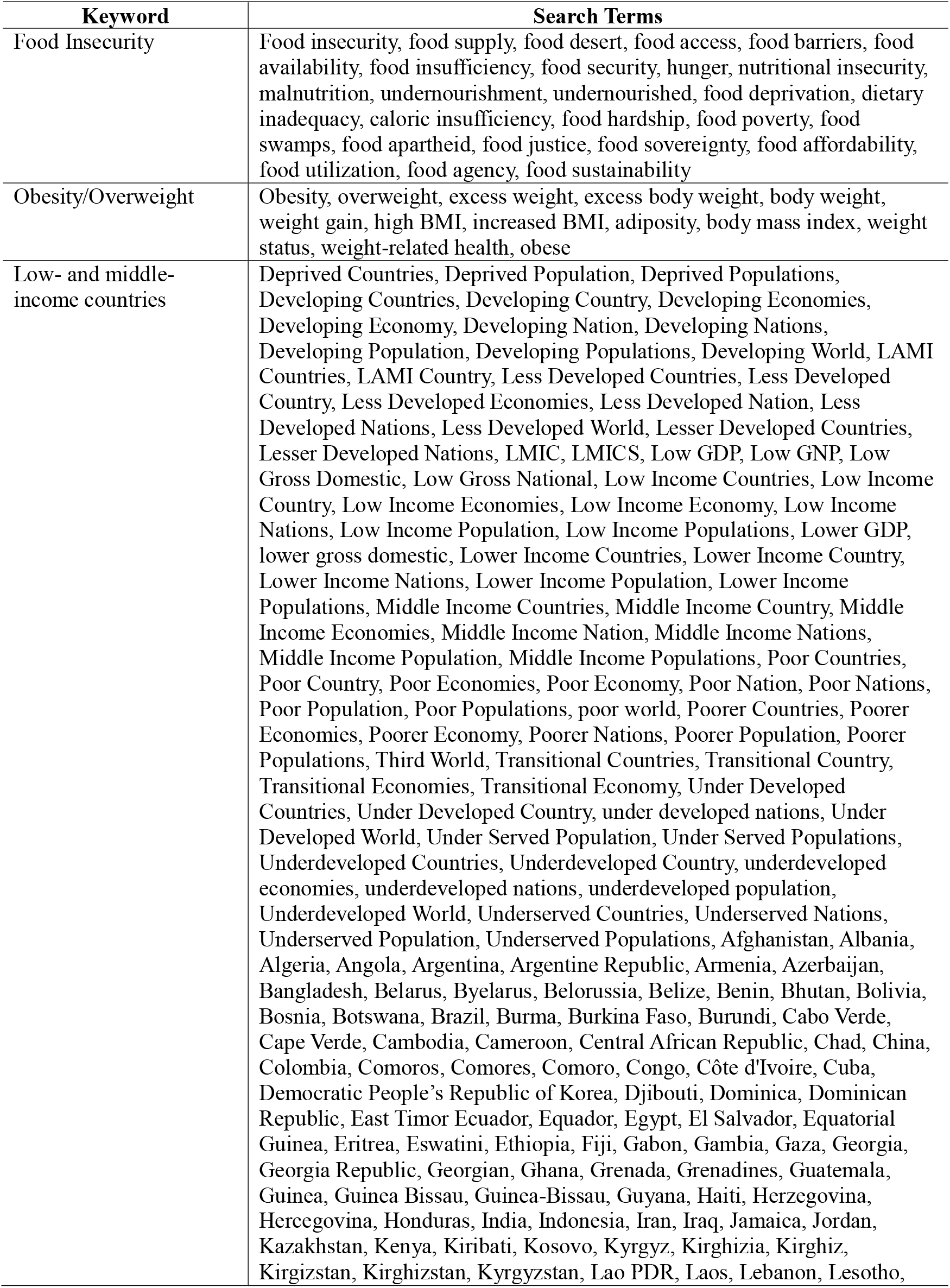
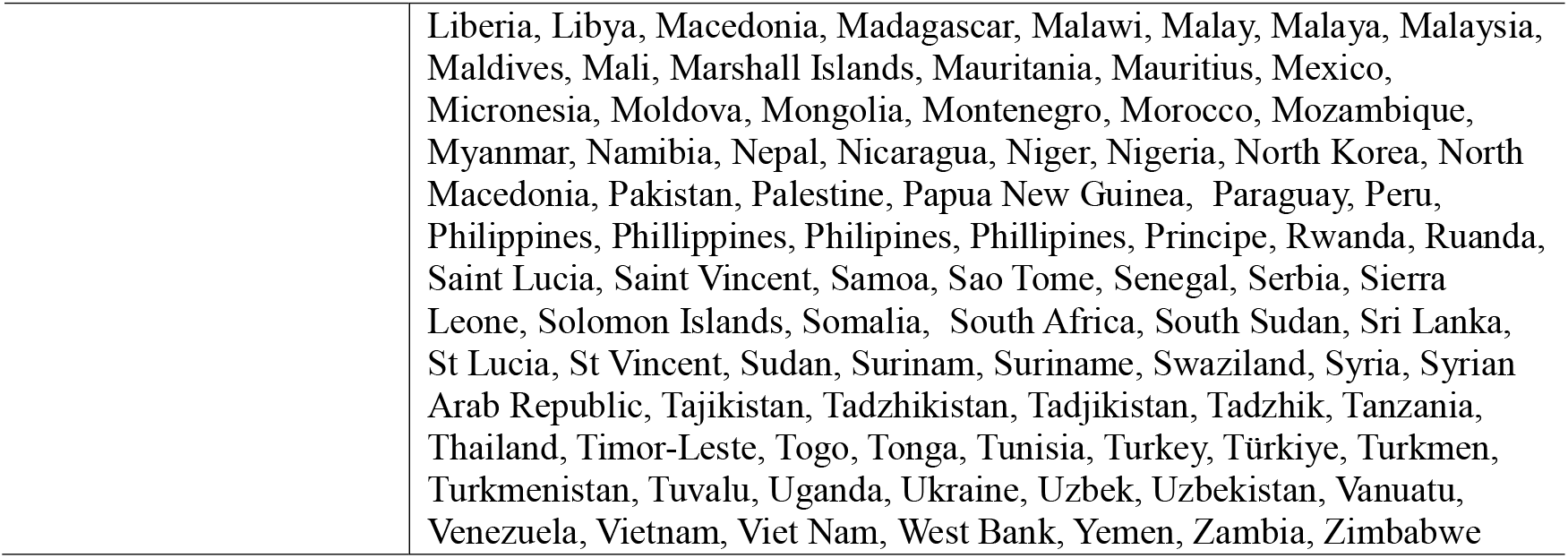
Search terms for all databases.

For grey literature sources, searches will be conducted directly in Google Scholar and ProQuest Dissertations & Theses Global using variations of the review’s title. Due to Google Scholar’s inconsistent and profuse number of search results, five combinations of keywords will be entered, sorting the results by relevancy. The first ten pages of results of each keyword combination (100 results per combination; 500 total) will be screened for relevant articles to maintain a reasonable amount of manual screening. The same combinations of keywords will be used for ProQuest Dissertations & Theses Global and screening the first five pages of results for each keyword combination (75 results per combination; 375 total). The search strings used for Google Scholar and ProQuest Dissertations & Theses Global are presented in **S4 and S5 Files**.

### Selection Process

All screening, duplication removal, and data extraction will be conducted using Covidence systematic review software (Veritas Health Innovation, Melbourne, Australia; available at www.covidence.org). Screening will be conducted in two stages: title and abstract review, followed by full-text review. Both stages will follow the predefined inclusion and exclusion criteria. Two reviewers will independently screen the records, with any disagreements resolved by discussion or adjudication by a third reviewer. For Spanish-language studies, full-text review will be conducted by one reviewer, with any unclear or borderline cases discussed with a second reviewer to reach consensus. The study selection process will be documented using a flow diagram developed in accordance with the PRISMA 2020 guidelines (31). This will detail the number of records identified, screened, excluded, and ultimately included at each stage of the review.

### Data collection process and data items

For all studies meeting the inclusion criteria, data will be extracted using a standardized and pre-piloted extraction form developed by the review team. Two reviewers will independently extract data, and any discrepancies will be resolved through discussion or by consulting a third reviewer. Where data are ambiguous or missing, we will not contact primary study authors and will rely solely on the information reported in the publication.

Extracted data will include the following:

- Study characteristics: author(s), year of publication, study period, country and region, country income classification, study design, sample size, participant demographics (e.g., age, gender), setting (urban or rural), and any reported vulnerability to extreme events (e.g., conflict, climate-related disasters).
- Exposure assessment: type of food insecurity measurement tool, level of measurement (household or individual), and categorization of food insecurity (e.g., mild, moderate, severe).
- Outcomes: methods used to assess body composition (e.g., BMI, waist circumference), source of anthropometric data (e.g., self-reported, measured, medical records), and outcome definitions (e.g., international or region-specific cutoffs). Both continuous and categorical assessments of BMI and WC will be included.
- Effect measures: estimates of association between food insecurity and overweight/obesity (e.g., odds ratios, relative risks), variables included in adjusted models, and reported confidence intervals.
- Additional data: proposed mechanisms linking food insecurity to overweight/obesity (when applicable), any potential sources of bias, any disclosure of funding sources, and conflicts of interest.

Publications derived from the same study will be identified and consolidated. A primary reference will be selected for reporting purposes. When related publications are suspected, we will compare author names, study locations, start dates and duration, and study sponsors or ethics approval numbers to confirm linkage.

For studies that explore potential mechanisms linking food insecurity with overweight/obesity, relevant qualitative or quantitative data that describes or tests explanatory pathways will be extracted. These may include narrative descriptions of hypothesized mechanisms or empirical assessments of intermediary factors (e.g., changes in diet quality, food purchasing patterns, or physical activity). Identified mechanisms will be described and grouped thematically. When possible, they will also be mapped onto existing conceptual frameworks.

The primary outcome of interest is overweight and/or obesity among adults experiencing food insecurity. Eligible studies must assess body composition using BMI, waist circumference, or both. Studies will be included regardless of whether they use internationally recognized cutoffs (e.g., BMI ≥25 for overweight and ≥30 for obesity) or region-specific thresholds, such as those applied in some Asian populations. Measures of central adiposity using WC will be accepted either alone or in combination with BMI. Both self-reported and instrument-based measures will be included, provided the methods are clearly described. When possible, results will be stratified by measurement type to account for potential differences in accuracy.

### Study Risk of Bias Assessment

Risk of bias for each included study will be assessed using the Joanna Briggs Institute (JBI) Critical Appraisal Tools, with the specific checklist applied based on the study design (i.e., cross-sectional, cohort, or case-control). Two reviewers will independently assess each study using the relevant tool. Any discrepancies will be resolved through discussion or, if needed, by a third reviewer. Risk of bias assessments will inform the interpretation of findings but will not be used as grounds for study exclusion.

### Data analysis and synthesis

The unit of analysis in this review will be the study population or subgroup as defined by each included study (e.g., adults, women). Studies will be grouped according to the direction and significance of the reported association between food insecurity and overweight/obesity. A positive association will be defined as a statistically significant direct relationship (i.e., food insecurity increases the risk of overweight or obesity), while a negative association will indicate a statistically significant inverse relationship (i.e., food insecurity is associated with a reduced risk). Studies reporting no association or mixed results—such as varying associations across subgroups defined by age, gender, or geographic location—will also be clearly categorized.

If sufficient data and methodological comparability exist across studies, a meta-analysis will be considered. The feasibility of conducting a meta-analysis will depend on the availability of effect measures, consistency in definitions and metrics, and the homogeneity of populations and study designs. If performed, heterogeneity will be assessed using Cochran’s Q test and I^2^ statistic. A fixed-or random-effects model will be chosen based on the degree of heterogeneity. We will use subgroup analyses to explore possible sources of heterogeneity, based on study-level characteristics such as gender, age group, region, and level of national income. Meta-regression may be performed to explore the influence of additional study-level covariates on the observed associations. If appropriate, sensitivity analyses will be conducted by excluding studies with high risk of bias or those using non-standard definitions of food insecurity or overweight/obesity. Forest plots will be used to display results, and publication bias will be assessed using funnel plots. All statistical analyses will be conducted using Stata (version 18.5; StataCorp LLC, College Station, TX, USA).

If a meta-analysis is not feasible due to substantial heterogeneity, we will present the results in a narrative synthesis to describe and interpret findings from the included studies. This synthesis will be supplemented by summary tables providing study characteristics and key findings related to the association between food insecurity and overweight or obesity. The quality of the evidence will be discussed based on the results of the risk of bias assessment. Where relevant, findings will be mapped against hypothesized mechanisms linking food insecurity to overweight/obesity, especially those described in Farrell et al. (2018), as well as any additional mechanisms identified during data extraction (23).

Given the observational nature of the studies and the likelihood of diverse populations, settings, and study designs, we do not plan to conduct a formal certainty of evidence assessment at this stage. Such assessments are most applicable when a meta-analysis is conducted. If a meta-analysis becomes feasible during the review process, we will consider incorporating an appropriate certainty assessment for key outcomes accordingly.

### Ethics and dissemination

This review will rely solely on data from previously published studies; therefore, ethical approval is not required. Any important amendments to the protocol will be clearly documented and updated in the PROSPERO record, as well as reported in the final publication. The findings of the systematic review will be shared through publication in a peer-reviewed journal and presented at relevant academic conferences to support evidence-based research and policy development.

## Discussion

This systematic review seeks to address a critical gap in the literature by synthesizing observational evidence on the association between food insecurity and overweight/obesity among adults in LMICs. Although much of the existing research and systematic reviews have focused on HICs, where food insecurity has been linked to obesity particularly among women, evidence from LMICs remains limited. Moradi et al. (2018) conducted a global systematic review and meta-analysis and found that in more economically developed countries, food insecurity was significantly associated with obesity, while in less developed countries, it was more strongly linked to underweight (29). The authors concluded that no clear association between food insecurity and overweight/obesity existed in developing countries. Similarly, a review by the Food and Agriculture Organization (FAO) highlighted that most positive associations between food insecurity and obesity were observed in women in HICs, while findings from LMICs were limited and inconclusive (30). Although these reviews did not identify a consistent relationship between food insecurity and overweight/obesity in LMICs, several individual studies within these settings did report significant associations. This suggests a need to focus more specifically on LMICs to better understand the mechanisms that may drive or mitigate the relationship between food insecurity and overweight or obesity status.

A key contribution of this review will be its focus not only on the direction and strength of the food insecurity–overweight/obesity associations, but also on the mechanisms and contextual factors that may influence such relationships. Farrell et al. (2018) proposed five key pathways through which food insecurity may contribute to obesity in LMICs: (1) the affordability and greater availability of energy-dense processed foods; (2) reduced dietary quantity and diversity; (3) spatial and temporal barriers to consistently and reliably accessing nutritious food (e.g., long distances from food sources or limited physical ability to travel); (4) intra-household food distribution inequalities; and (5) non-dietary mechanisms, such as reduced physical activity or stress-related behaviors (23). These pathways are not mutually exclusive and may operate differently depending on gender, age, income level, urbanicity, and national context. Authors also emphasized that the heterogeneity in existing studies may be due to unmeasured mediators and contextual interactions that are not yet fully understood, such as length of time a household is exposed to food insecurity or social and cultural acceptability (23). By including more recent studies and systematically documenting proposed mechanisms and contextual modifiers, this review aims to deepen the understanding of how and why food insecurity may be linked to overweight/obesity in diverse LMIC settings. The present review also aims to examine whether new or underexplored pathways have emerged or are becoming hypothesized in recent years.

The findings of this review have direct implications for health research and policy in LMICs, many of which are facing a growing burden of diet-related noncommunicable diseases. As several LMICs (e.g., Mexico, Colombia, Peru, Argentina) implement front-of-package warning labels, sugary drink taxes, and other food environment policies to curb obesity, understanding how these interventions interact with food insecurity is essential (40). If food-insecure populations are disproportionately exposed to unhealthy food environments or unable to benefit from policy-driven improvements, interventions may need to be adapted to ensure equity and effectiveness. Insights from this review can help inform the design of nutrition and social protection policies that more comprehensively address the triple burden of malnutrition and support healthy food access for the most vulnerable populations.

This review has several strengths. It will employ a systematic and transparent approach following PRISMA guidelines, with predefined eligibility criteria and a rigorous multi-stage screening and data extraction process. In addition to mapping the evidence on the association between food insecurity and overweight/obesity in LMICs, the review will go further by identifying and summarizing proposed mechanisms and contextual factors, offering valuable insights for both researchers and policymakers. Importantly, unlike previous reviews that limited inclusion to English-language publications, this review will also incorporate studies and evaluations published in Spanish—a key addition given that Spanish is the primary language in several LMICs (33). Nonetheless, some limitations must be acknowledged. First, variations in how food insecurity is measured across studies may limit comparability. Second, BMI cutoffs for defining overweight and obesity may differ by country or region, which could affect outcome classification and synthesis. Lastly, despite a comprehensive search strategy put in place, there may be limited data available from low-income countries, potentially underrepresenting the most vulnerable settings.

## Data Availability

This study is a protocol for a systematic review and does not involve the generation or analysis of primary data. All data used will be obtained from previously published studies.

## Publication Status

This protocol has been developed for the ongoing systematic review titled “The association between food insecurity and overweight/obesity among adults in low- and middle-income countries: A systematic review of observational studies”. Any modifications made to the protocol during the course of the review will be documented and updated in the PROSPERO registration database (Registration number: CRD420251076653). PROSPERO is an international prospective register of systematic reviews designed to promote transparency and reduce the risk of duplication. These changes will also be reported in the published systematic review.

## Funding source

This study is supported by Bloomberg Philanthropies. The funder had no role in the development of the protocol, including the study design, data extraction strategy, analysis plan, or decision to submit for publication.

## Notes

### Competing Interest Statement

The authors have declared no competing interest.

