## Supplemental Material for "Exploring the association between food insecurity and overweight/obesity among adults in low- and middle-income countries: A systematic review protocol"

**S1. PRISMA-P Checklist**

PRISMA-P (Preferred Reporting Items for Systematic review and Meta-Analysis Protocols) 2015 checklist: recommended items to address in a systematic review protocol

| Section and topic | Item No | Checklist item | Page number |
| --- | --- | --- | --- |
| ADMINISTRATIVE INFORMATION | | |  |
| Title: |  |  |  |
| Identification | 1a | Identify the report as a protocol of a systematic review | 1 |
| Update | 1b | If the protocol is for an update of a previous systematic review, identify as such |  |
| Registration | 2 | If registered, provide the name of the registry (such as PROSPERO) and registration number | 2 |
| Authors: |  |  |  |
| Contact | 3a | Provide name, institutional affiliation, e-mail address of all protocol authors; provide physical mailing address of corresponding author | 1 |
| Contributions | 3b | Describe contributions of protocol authors and identify the guarantor of the review |  |
| Amendments | 4 | If the protocol represents an amendment of a previously completed or published protocol, identify as such and list changes; otherwise, state plan for documenting important protocol amendments | 11 |
| Support: |  |  |  |
| Sources | 5a | Indicate sources of financial or other support for the review |  |
| Sponsor | 5b | Provide name for the review funder and/or sponsor | 13 |
| Role of sponsor or funder | 5c | Describe roles of funder(s), sponsor(s), and/or institution(s), if any, in developing the protocol |  |
| INTRODUCTION | | |  |
| Rationale | 6 | Describe the rationale for the review in the context of what is already known | 3-4 |
| Objectives | 7 | Provide an explicit statement of the question(s) the review will address with reference to participants, interventions, comparators, and outcomes (PICO) | 4 |
| METHODS | | |  |
| Eligibility criteria | 8 | Specify the study characteristics (such as PICO, study design, setting, time frame) and report characteristics (such as years considered, language, publication status) to be used as criteria for eligibility for the review | 4-6 |
| Information sources | 9 | Describe all intended information sources (such as electronic databases, contact with study authors, trial registers or other grey literature sources) with planned dates of coverage | 6 |
| Search strategy | 10 | Present draft of search strategy to be used for at least one electronic database, including planned limits, such that it could be repeated | 7-8 |
| Study records: |  |  | 8-9 |
| Data management | 11a | Describe the mechanism(s) that will be used to manage records and data throughout the review |  |
| Selection process | 11b | State the process that will be used for selecting studies (such as two independent reviewers) through each phase of the review (that is, screening, eligibility and inclusion in meta-analysis) | 8-9 |
| Data collection process | 11c | Describe planned method of extracting data from reports (such as piloting forms, done independently, in duplicate), any processes for obtaining and confirming data from investigators | 8-9 |
| Data items | 12 | List and define all variables for which data will be sought (such as PICO items, funding sources), any pre-planned data assumptions and simplifications | 8-9 |
| Outcomes and prioritization | 13 | List and define all outcomes for which data will be sought, including prioritization of main and additional outcomes, with rationale | 9 |
| Risk of bias in individual studies | 14 | Describe anticipated methods for assessing risk of bias of individual studies, including whether this will be done at the outcome or study level, or both; state how this information will be used in data synthesis | 9 |
| Data synthesis | 15a | Describe criteria under which study data will be quantitatively synthesised | 10 |
|  | 15b | If data are appropriate for quantitative synthesis, describe planned summary measures, methods of handling data and methods of combining data from studies, including any planned exploration of consistency (such as I^2^, Kendall’s τ) | 10 |
|  | 15c | Describe any proposed additional analyses (such as sensitivity or subgroup analyses, meta-regression) | 10 |
|  | 15d | If quantitative synthesis is not appropriate, describe the type of summary planned | 10 |
| Meta-bias(es) | 16 | Specify any planned assessment of meta-bias(es) (such as publication bias across studies, selective reporting within studies) | 10 |
| Confidence in cumulative evidence | 17 | Describe how the strength of the body of evidence will be assessed (such as GRADE) | 10 |

**S2. Search strategy (English)**

A language filter will be applied to include articles in English and Spanish across the following databases: PubMed, EMBASE, Global Health, and Scopus. For LILACS, the search strategy was translated into Spanish, as many articles in this database do not provide titles or abstracts in English—unlike the other databases listed above.

**PUBMED**

("Food Insecurity"[Mesh] OR “Food supply”[MeSH] OR “food supply”[tiab] OR “food desert”[tiab] OR “food deserts”[tiab] OR "food insecurity"[tiab] OR "food access"[tiab:~2] OR “food barriers”[tiab:~2] OR "food availability"[tiab] OR "food insufficiency"[tiab] OR "food security"[tiab:~2] OR hunger[tiab] OR "nutritional insecurity"[tiab] OR malnutrition[tiab] OR undernourishment[tiab] OR "food deprivation"[tiab] OR "dietary inadequacy"[tiab] OR "caloric insufficiency"[tiab] OR "food hardship"[tiab] OR "food poverty"[tiab] OR "food swamps"[tiab] OR "food justice"[tiab] OR "food access disparities"[tiab] OR starvation[tiab] OR “food apartheid”[tiab] OR undernourished[tiab] OR “food sovereignty”[tiab] OR “food affordability”[tiab] OR “food utilization”[tiab] OR “food agency”[tiab] OR “food sustainability”[tiab])

AND

("Obesity"[Mesh] OR "Overweight"[Mesh] OR obesity[tiab] OR overweight[tiab] OR "excess weight"[tiab] OR "excess body weight"[tiab] OR "body weight"[tiab] OR "weight gain"[tiab] OR "high BMI"[tiab] OR "increased BMI"[tiab] OR adiposity[tiab] OR "body mass index"[tiab] OR "weight status"[tiab] OR "weight-related health"[tiab] OR obese[tiab])

AND

(Deprived Countries[tw] OR Deprived Population[tw] OR Deprived Populations[tw] OR Developing Countries[tw] OR Developing Country[tw] OR Developing Economies[tw] OR Developing Economy[tw] OR Developing Nation[tw] OR Developing Nations[tw] OR Developing Population[tw] OR Developing Populations[tw] OR Developing World[tw] “Global South”[tw]OR LAMI Countries[tw] OR LAMI Country[tw] OR Less Developed Countries[tw] OR Less Developed Country[tw] OR Less Developed Economies[tw] OR Less Developed Nation[tw] OR Less Developed Nations[tw] OR Less Developed World[tw] OR Lesser Developed Countries[tw] OR Lesser Developed Nations[tw] OR LMIC[tw] OR LMICS[tw] OR Low GDP[tw] OR Low GNP[tw] OR Low Gross Domestic[tw] OR Low Gross National[tw] OR Low Income Countries[tw] OR Low Income Country[tw] OR Low Income Economies[tw] OR Low Income Economy[tw] OR Low Income Nations[tw] OR Low Income Population[tw] OR Low Income Populations[tw] OR Lower GDP[tw] OR lower gross domestic[tw] OR Lower Income Countries[tw] OR Lower Income Country[tw] OR Lower Income Nations[tw] OR Lower Income Population[tw] OR Lower Income Populations[tw] OR Middle Income Countries[tw] OR Middle Income Country[tw] OR Middle Income Economies[tw] OR Middle Income Nation[tw] OR Middle Income Nations[tw] OR Middle Income Population[tw] OR Middle Income Populations[tw] OR Poor Countries[tw] OR Poor Country[tw] OR Poor Economies[tw] OR Poor Economy[tw] OR Poor Nation[tw] OR Poor Nations[tw] OR Poor Population[tw] OR Poor Populations[tw] OR poor world[tw] OR Poorer Countries[tw] OR Poorer Economies[tw] OR Poorer Economy[tw] OR Poorer Nations[tw] OR Poorer Population[tw] OR Poorer Populations[tw] OR Third World[tw] OR Transitional Countries[tw] OR Transitional Country[tw] OR Transitional Economies[tw] OR Transitional Economy[tw] OR Under Developed Countries[tw] OR Under Developed Country[tw] OR under developed nations[tw] OR Under Developed World[tw] OR Under Served Population[tw] OR Under Served Populations[tw] OR Underdeveloped Countries[tw] OR Underdeveloped Country[tw] OR underdeveloped economies[tw] OR underdeveloped nations[tw] OR underdeveloped population[tw] OR Underdeveloped World[tw] OR Underserved Countries[tw] OR Underserved Nations[tw] OR Underserved Population[tw] OR Underserved Populations[tw] OR Afghanistan[tw] OR Albania[tw] OR Algeria[tw] OR Angola[tw] OR Argentina[tw] OR “Argentine Republic”[tw] OR Armenia[tw] OR Azerbaijan[tw] OR Bangladesh[tw] OR Belarus[tw] OR Byelarus[tw] OR Belorussia[tw] OR Belize[tw] OR Benin[tw] OR Bhutan[tw] OR Bolivia[tw] OR Bosnia[tw] OR Botswana[tw] OR Brazil[tw] OR Burma[tw] OR “Burkina Faso”[tw] OR Burundi[tw] OR “Cabo Verde”[tw] OR “Cape Verde”[tw] OR Cambodia[tw] OR Cameroon[tw] OR “Central African Republic”[tw] OR Chad[tw] OR China[tw] OR Colombia[tw] OR Comoros[tw] OR Comores[tw] OR Comoro[tw] OR Congo[tw] OR “Côte d'Ivoire”[tw] OR Cuba[tw] OR “Democratic People’s Republic of Korea”[tw] OR Djibouti[tw] OR Dominica[tw] OR “Dominican Republic”[tw] OR “East Timor”[tw] OR Ecuador[tw] OR Equador[tw] OR Egypt[tw] OR “El Salvador”[tw] OR Eritrea[tw] OR Eswatini[tw] OR Ethiopia[tw] OR Fiji[tw] OR Gabon[tw] OR Gambia[tw] OR Gaza[tw] OR Georgia[tw] OR “Georgia Republic”[tw] OR Georgian[tw] OR Ghana[tw] OR Grenada[tw] OR Grenadines[tw] OR Guatemala[tw] OR Guinea[tw] OR “Guinea Bissau”[tw] OR Guinea-Bissau[tw] OR Guyana[tw] OR Haiti[tw] OR Herzegovina[tw] OR Hercegovina[tw] OR Honduras[tw] OR India[tw] OR Indonesia[tw] OR Iran[tw] OR Iraq[tw] OR Jamaica[tw] OR Jordan[tw] OR Kazakhstan[tw] OR Kenya[tw] OR Kiribati[tw] OR Kosovo[tw] OR Kyrgyz[tw] OR Kirghizia[tw] OR Kirghiz[tw] OR Kirgizstan[tw] OR Kirghizstan[tw] OR Kyrgyzstan[tw] OR “Lao PDR”[tw] OR Laos[tw] OR Lebanon[tw] OR Lesotho[tw] OR Liberia[tw] OR Libya[tw] OR Macedonia[tw] OR Madagascar[tw] OR Malawi[tw] OR Malay[tw] OR Malaya[tw] OR Malaysia[tw] OR Maldives[tw] OR Mali[tw] OR “Marshall Islands”[tw] OR Mauritania[tw] OR Mauritius[tw] OR Mexico[tw] OR Micronesia[tw] OR Moldova[tw] OR Mongolia[tw] OR Montenegro[tw] OR Morocco[tw] OR Mozambique[tw] OR Myanmar[tw] OR Namibia[tw] OR Nepal[tw] OR Nicaragua[tw] OR Niger[tw] OR Nigeria[tw] OR “North Korea”[tw] OR Pakistan[tw] OR Palestine[tw] OR Panama[tw] OR “Papua New Guinea”[tw] OR Paraguay[tw] OR Peru[tw] OR Philippines[tw] OR Phillippines[tw] OR Philipines[tw] OR Phillipines[tw] OR Principe[tw] OR Rwanda[tw] OR Ruanda[tw] OR “Saint Lucia”[tw] OR “Saint Vincent”[tw] OR Samoa[tw] OR “Sao Tome”[tw] OR Senegal[tw] OR Serbia[tw] OR “Sierra Leone”[tw] OR “Solomon Islands”[tw] OR Somalia[tw] OR “South Africa”[tw] OR “South Sudan”[tw] OR “Sri Lanka”[tw] OR “St Lucia”[tw] OR “St Vincent”[tw] OR Sudan[tw] OR Surinam[tw] OR Suriname[tw] OR Swaziland[tw] OR Syria[tw] OR “Syrian Arab Republic”[tw] OR Tajikistan[tw] OR Tadzhikistan[tw] OR Tadjikistan[tw] OR Tadzhik[tw] OR Tanzania[tw] OR Thailand[tw] OR Timor-Leste[tw] OR Togo[tw] OR Tonga[tw] OR Tunisia[tw] OR Turkey[tw] OR Türkiye[tw] OR Turkmen[tw] OR Turkmenistan[tw] OR Tuvalu[tw] OR Uganda[tw] OR Ukraine[tw] OR Uzbek[tw] OR Uzbekistan[tw] OR Vanuatu[tw] OR Venezuela[tw] OR Vietnam[tw] OR “Viet Nam”[tw] OR “West Bank”[tw] OR Yemen[tw] OR Zambia[tw] OR Zimbabwe[tw])

AND

("2000/01/01"[PDAT] : "3000"[PDAT])

**EMBASE**

("Food Insecurity"/exp OR “Food availability”/exp OR “food supply”:ab,ti OR “food desert”:ab,ti OR “food deserts”:ab,ti OR "food insecurity":ab,ti OR (food NEAR/2 access):ab,ti OR (food NEAR/2 barriers):ab,ti OR "food availability":ab,ti OR "food insufficiency":ab,ti OR (food NEAR/2 security):ab,ti OR hunger:ab,ti OR "nutritional insecurity":ab,ti OR malnutrition:ab,ti OR undernourishment:ab,ti OR "food deprivation":ab,ti OR "dietary inadequacy":ab,ti OR "caloric insufficiency":ab,ti OR "food hardship":ab,ti OR "food poverty":ab,ti OR "food swamps":ab,ti OR "food justice":ab,ti OR "food access disparities":ab,ti OR starvation:ab,ti OR “food apartheid”:ab,ti OR undernourished:ab,ti OR “food sovereignty”:ab,ti OR “food affordability”:ab,ti OR “food utilization”:ab,ti OR “food agency”:ab,ti OR “food sustainability”:ab,ti)

AND

("Obesity"/exp OR obesity:ab,ti OR overweight:ab,ti OR "excess weight":ab,ti OR "excess body weight":ab,ti OR "body weight":ab,ti OR "weight gain":ab,ti OR "high BMI":ab,ti OR "increased BMI":ab,ti OR adiposity:ab,ti OR "body mass index":ab,ti OR "weight status":ab,ti OR "weight-related health":ab,ti OR obese:ab,ti)

AND

(“Deprived Countries”:ti,ab,kw OR “Deprived Population”:ti,ab,kw OR “Deprived Populations”:ti,ab,kw OR “Developing Countries”:ti,ab,kw OR “Developing Country”:ti,ab,kw OR “Developing Economies”:ti,ab,kw OR “Developing Economy”:ti,ab,kw OR “Developing Nation”:ti,ab,kw OR “Developing Nations”:ti,ab,kw OR “Developing Population”:ti,ab,kw OR “Developing Populations”:ti,ab,kw OR “Developing World”:ti,ab,kw “Global South”:ti,ab,kw OR “LAMI Countries”:ti,ab,kw OR “LAMI Country”:ti,ab,kw OR “Less Developed Countries”:ti,ab,kw OR “Less Developed Country”:ti,ab,kw OR “Less Developed Economies”:ti,ab,kw “Global South”:ti,ab,kw OR “Less Developed Nation”:ti,ab,kw OR “Less Developed Nations”:ti,ab,kw OR “Less Developed World”:ti,ab,kw OR “Lesser Developed Countries”:ti,ab,kw OR “Lesser Developed Nations”:ti,ab,kw OR LMIC:ti,ab,kw OR LMICS:ti,ab,kw OR “Low GDP”:ti,ab,kw OR “Low GNP”:ti,ab,kw OR “Low Gross Domestic”:ti,ab,kw OR “Low Gross National”:ti,ab,kw OR “Low Income Countries”:ti,ab,kw OR “Low Income Country”:ti,ab,kw OR “Low Income Economies”:ti,ab,kw OR “Low Income Economy”:ti,ab,kw OR “Low Income Nations”:ti,ab,kw OR “Low Income Population”:ti,ab,kw OR “Low Income Populations”:ti,ab,kw OR “Lower GDP”:ti,ab,kw OR “lower gross domestic”:ti,ab,kw OR “Lower Income Countries”:ti,ab,kw OR “Lower Income Country”:ti,ab,kw OR “Lower Income Nations”:ti,ab,kw OR “Lower Income Population”:ti,ab,kw OR “Lower Income Populations”:ti,ab,kw OR “Middle Income Countries”:ti,ab,kw OR “Middle Income Country”:ti,ab,kw OR “Middle Income Economies”:ti,ab,kw OR “Middle Income Nation”:ti,ab,kw OR “Middle Income Nations”:ti,ab,kw OR “Middle Income Population”:ti,ab,kw OR “Middle Income Populations”:ti,ab,kw OR “Poor Countries”:ti,ab,kw OR “Poor Country”:ti,ab,kw OR “Poor Economies”:ti,ab,kw OR “Poor Economy”:ti,ab,kw OR “Poor Nation”:ti,ab,kw OR “Poor Nations”:ti,ab,kw OR “Poor Population”:ti,ab,kw OR “Poor Populations”:ti,ab,kw OR “poor world”:ti,ab,kw OR “Poorer Countries”:ti,ab,kw OR “Poorer Economies”:ti,ab,kw OR “Poorer Economy”:ti,ab,kw OR “Poorer Nations”:ti,ab,kw OR “Poorer Population”:ti,ab,kw OR “Poorer Populations”:ti,ab,kw OR “Third World”:ti,ab,kw OR “Transitional Countries”:ti,ab,kw OR “Transitional Country”:ti,ab,kw OR “Transitional Economies”:ti,ab,kw OR “Transitional Economy”:ti,ab,kw OR “Under Developed Countries”:ti,ab,kw OR “Under Developed Country”:ti,ab,kw OR “under developed nations”:ti,ab,kw OR “Under Developed World”:ti,ab,kw OR “Under Served Population”:ti,ab,kw OR “Under Served Populations”:ti,ab,kw OR “Underdeveloped Countries”:ti,ab,kw OR “Underdeveloped Country”:ti,ab,kw OR “underdeveloped economies”:ti,ab,kw OR “underdeveloped nations”:ti,ab,kw OR “underdeveloped population”:ti,ab,kw OR “Underdeveloped World”:ti,ab,kw OR “Underserved Countries”:ti,ab,kw OR “Underserved Nations”:ti,ab,kw OR “Underserved Population”:ti,ab,kw OR “Underserved Populations”:ti,ab,kw OR Afghanistan:ti,ab,kw OR Albania:ti,ab,kw OR Algeria:ti,ab,kw OR Angola:ti,ab,kw OR Argentina:ti,ab,kw OR “Argentine Republic”:ti,ab,kw OR Armenia:ti,ab,kw OR Azerbaijan:ti,ab,kw OR Bangladesh:ti,ab,kw OR Belarus:ti,ab,kw OR Byelarus:ti,ab,kw OR Belorussia:ti,ab,kw OR Belize:ti,ab,kw OR Benin:ti,ab,kw OR Bhutan:ti,ab,kw OR Bolivia:ti,ab,kw OR Bosnia:ti,ab,kw OR Botswana:ti,ab,kw OR Brazil:ti,ab,kw OR Burma:ti,ab,kw OR “Burkina Faso”:ti,ab,kw OR Burundi:ti,ab,kw OR “Cabo Verde”:ti,ab,kw OR “Cape verde”:ti,ab,kw OR Cambodia:ti,ab,kw OR Cameroon:ti,ab,kw OR “Central African Republic”:ti,ab,kw OR Chad:ti,ab,kw OR China:ti,ab,kw OR Colombia:ti,ab,kw OR Comoros:ti,ab,kw OR Comores:ti,ab,kw OR Comoro:ti,ab,kw OR Congo:ti,ab,kw OR “Côte dIvoire”:ti,ab,kw OR Cuba:ti,ab,kw OR “Democratic Peoples Republic of Korea”:ti,ab,kw OR Djibouti:ti,ab,kw OR Dominica:ti,ab,kw OR “Dominican Republic”:ti,ab,kw OR “East Timor”:ti,ab,kw OR Ecuador:ti,ab,kw OR Equador:ti,ab,kw OR Egypt:ti,ab,kw OR “El Salvador”:ti,ab,kw OR Eritrea:ti,ab,kw OR Eswatini:ti,ab,kw OR Ethiopia:ti,ab,kw OR Fiji:ti,ab,kw OR Gabon:ti,ab,kw OR Gambia:ti,ab,kw OR Gaza:ti,ab,kw OR Georgia:ti,ab,kw OR “Georgia Republic”:ti,ab,kw OR Georgian:ti,ab,kw OR Ghana:ti,ab,kw OR Grenada:ti,ab,kw OR Grenadines:ti,ab,kw OR Guatemala:ti,ab,kw OR Guinea:ti,ab,kw OR “Guinea Bissau”:ti,ab,kw OR Guinea-Bissau:ti,ab,kw OR Guyana:ti,ab,kw OR Haiti:ti,ab,kw OR Herzegovina:ti,ab,kw OR Hercegovina:ti,ab,kw OR Honduras:ti,ab,kw OR India:ti,ab,kw OR Indonesia:ti,ab,kw OR Iran:ti,ab,kw OR Iraq:ti,ab,kw OR Jamaica:ti,ab,kw OR Jordan:ti,ab,kw OR Kazakhstan:ti,ab,kw OR Kenya:ti,ab,kw OR Kiribati:ti,ab,kw OR Kosovo:ti,ab,kw OR Kyrgyz:ti,ab,kw OR Kirghizia:ti,ab,kw OR Kirghiz:ti,ab,kw OR Kirgizstan:ti,ab,kw OR Kirghizstan:ti,ab,kw OR Kyrgyzstan:ti,ab,kw OR “Lao PDR”:ti,ab,kw OR Laos:ti,ab,kw OR Lebanon:ti,ab,kw OR Lesotho:ti,ab,kw OR Liberia:ti,ab,kw OR Libya:ti,ab,kw OR Macedonia:ti,ab,kw OR Madagascar:ti,ab,kw OR Malawi:ti,ab,kw OR Malay:ti,ab,kw OR Malaya:ti,ab,kw OR Malaysia:ti,ab,kw OR Maldives:ti,ab,kw OR Mali:ti,ab,kw OR “Marshall Islands”:ti,ab,kw OR Mauritania:ti,ab,kw OR Mauritius:ti,ab,kw OR Mexico:ti,ab,kw OR Micronesia:ti,ab,kw OR Moldova:ti,ab,kw OR Mongolia:ti,ab,kw OR Montenegro:ti,ab,kw OR Morocco:ti,ab,kw OR Mozambique:ti,ab,kw OR Myanmar:ti,ab,kw OR Namibia:ti,ab,kw OR Nepal:ti,ab,kw OR Nicaragua:ti,ab,kw OR Niger:ti,ab,kw OR Nigeria:ti,ab,kw OR “North Korea”:ti,ab,kw OR Pakistan:ti,ab,kw OR Palestine:ti,ab,kw OR Panama:ti,ab,kw OR “Papua New Guinea”:ti,ab,kw OR Paraguay:ti,ab,kw OR Peru:ti,ab,kw OR Philippines:ti,ab,kw OR Phillippines:ti,ab,kw OR Philipines:ti,ab,kw OR Phillipines:ti,ab,kw OR Principe:ti,ab,kw OR Rwanda:ti,ab,kw OR Ruanda:ti,ab,kw OR “Saint Lucia”:ti,ab,kw OR “Saint Vincent”:ti,ab,kw OR Samoa:ti,ab,kw OR “Sao Tome”:ti,ab,kw OR Senegal:ti,ab,kw OR Serbia:ti,ab,kw OR “Sierra Leone”:ti,ab,kw OR “Solomon Islands”:ti,ab,kw OR Somalia:ti,ab,kw OR “South Africa”:ti,ab,kw OR “South Sudan”:ti,ab,kw OR “Sri Lanka”:ti,ab,kw OR “St Lucia”:ti,ab,kw OR “St Vincent”:ti,ab,kw OR Sudan:ti,ab,kw OR Surinam:ti,ab,kw OR Suriname:ti,ab,kw OR Swaziland:ti,ab,kw OR Syria:ti,ab,kw OR “Syrian Arab Republic”:ti,ab,kw OR Tajikistan:ti,ab,kw OR Tadzhikistan:ti,ab,kw OR Tadjikistan:ti,ab,kw OR Tadzhik:ti,ab,kw OR Tanzania:ti,ab,kw OR Thailand:ti,ab,kw OR Timor-Leste:ti,ab,kw OR Togo:ti,ab,kw OR Tonga:ti,ab,kw OR Tunisia:ti,ab,kw OR Turkey:ti,ab,kw OR Türkiye:ti,ab,kw OR Turkmen:ti,ab,kw OR Turkmenistan:ti,ab,kw OR Tuvalu:ti,ab,kw OR Uganda:ti,ab,kw OR Ukraine:ti,ab,kw OR Uzbek:ti,ab,kw OR Uzbekistan:ti,ab,kw OR Vanuatu:ti,ab,kw OR Venezuela:ti,ab,kw OR Vietnam:ti,ab,kw OR “Viet Nam”:ti,ab,kw OR “West Bank”:ti,ab,kw OR Yemen:ti,ab,kw OR Zambia:ti,ab,kw OR Zimbabwe:ti,ab,kw)

AND

[2000-2025]/py

**GLOBAL HEALTH**

(SU ("Food security" OR “Food supply”) OR AB (“food supply” OR “food desert” OR “food deserts” OR "food insecurity" OR food N2 access OR food N2 barriers OR "food availability" OR "food insufficiency" OR food N2 security OR hunger OR "nutritional insecurity" OR malnutrition OR undernourishment OR "food deprivation" OR "dietary inadequacy" OR "caloric insufficiency" OR "food hardship" OR "food poverty" OR "food swamps" OR "food justice" OR "food access disparities" OR starvation OR “food apartheid” OR undernourished OR “food sovereignty” OR “food affordability” OR “food utilization” OR “food agency” OR “food sustainability”) OR TI (“food supply” OR “food desert” OR “food deserts” OR "food insecurity" OR food N2 access OR food N2 barriers OR "food availability" OR "food insufficiency" OR food N2 security OR hunger OR "nutritional insecurity" OR malnutrition OR undernourishment OR "food deprivation" OR "dietary inadequacy" OR "caloric insufficiency" OR "food hardship" OR "food poverty" OR "food swamps" OR "food justice" OR "food access disparities" OR starvation OR “food apartheid” OR undernourished OR “food sovereignty” OR “food affordability” OR “food utilization” OR “food agency” OR “food sustainability”))

AND

(SU (Obesity OR Overweight) OR TI (obesity OR overweight OR "excess weight" OR "excess body weight" OR "body weight" OR "weight gain" OR "high BMI" OR "increased BMI" OR adiposity OR "body mass index" OR "weight status" OR "weight-related health" OR obese) OR AB (obesity OR overweight OR "excess weight" OR "excess body weight" OR "body weight" OR "weight gain" OR "high BMI" OR "increased BMI" OR adiposity OR "body mass index" OR "weight status" OR "weight-related health" OR obese))

AND

(AB (“Deprived Countries” OR “Deprived Population” OR “Deprived Populations” OR “Developing Countries” OR “Developing Country” OR “Developing Economies” OR “Developing Economy” OR “Developing Nation” OR “Developing Nations” OR “Developing Population” OR “Developing Populations” OR “Developing World” OR “Global South” OR “LAMI Countries” OR “LAMI Country” OR “Less Developed Countries” OR “Less Developed Country” OR “Less Developed Economies” OR “Less Developed Nation” OR “Less Developed Nations” OR “Less Developed World” OR “Lesser Developed Countries” OR “Lesser Developed Nations” OR LMIC OR LMICS OR “Low GDP” OR “Low GNP” OR “Low Gross Domestic” OR “Low Gross National” OR “Low Income Countries” OR “Low Income Country” OR “Low Income Economies” OR “Low Income Economy” OR “Low Income Nations” OR “Low Income Population” OR “Low Income Populations” OR “Lower GDP” OR “lower gross domestic” OR “Lower Income Countries” OR “Lower Income Country” OR “Lower Income Nations” OR “Lower Income Population” OR “Lower Income Populations” OR “Middle Income Countries” OR “Middle Income Country” OR “Middle Income Economies” OR “Middle Income Nation” OR “Middle Income Nations” OR “Middle Income Population” OR “Middle Income Populations” OR “Poor Countries” OR “Poor Country” OR “Poor Economies” OR “Poor Economy” OR “Poor Nation” OR “Poor Nations” OR “Poor Population” OR “Poor Populations” OR “poor world” OR “Poorer Countries” OR “Poorer Economies” OR “Poorer Economy” OR “Poorer Nations” OR “Poorer Population” OR “Poorer Populations” OR “Third World” OR “Transitional Countries” OR “Transitional Country” OR “Transitional Economies” OR “Transitional Economy” OR “Under Developed Countries” OR “Under Developed Country” OR “under developed nations” OR “Under Developed World” OR “Under Served Population” OR “Under Served Populations” OR “Underdeveloped Countries” OR “Underdeveloped Country” OR “underdeveloped economies” OR “underdeveloped nations” OR “underdeveloped population” OR “Underdeveloped World” OR “Underserved Countries” OR “Underserved Nations” OR “Underserved Population” OR “Underserved Populations” OR Afghanistan OR Albania OR Algeria OR Angola OR Argentina OR “Argentine Republic” OR Armenia OR Azerbaijan OR Bangladesh OR Belarus OR Byelarus OR Belorussia OR Belize OR Benin OR Bhutan OR Bolivia OR Bosnia OR Botswana OR Brazil OR Burma OR “Burkina Faso” OR Burundi OR “Cabo Verde” OR “Cape Verde” OR Cambodia OR Cameroon OR “Central African Republic” OR Chad OR China OR Colombia OR Comoros OR Comores OR Comoro OR Congo OR “Côte d’Ivoire” OR Cuba OR “Democratic Peoples Republic of Korea” OR Djibouti OR Dominica OR “Dominican Republic” OR “East Timor” OR Ecuador OR Equador OR Egypt OR “El Salvador” OR Eritrea OR Eswatini OR Ethiopia OR Fiji OR Gabon OR Gambia OR Gaza OR Georgia OR “Georgia Republic” OR Georgian OR Ghana OR Grenada OR Grenadines OR Guatemala OR Guinea OR “Guinea Bissau” OR Guinea-Bissau OR Guyana OR Haiti OR Herzegovina OR Hercegovina OR Honduras OR India OR Indonesia OR Iran OR Iraq OR Jamaica OR Jordan OR Kazakhstan OR Kenya OR Kiribati OR Kosovo OR Kyrgyz OR Kirghizia OR Kirghiz OR Kirgizstan OR Kirghizstan OR Kyrgyzstan OR “Lao PDR” OR Laos OR Lebanon OR Lesotho OR Liberia OR Libya OR Macedonia OR Madagascar OR Malawi OR Malay OR Malaya OR Malaysia OR Maldives OR Mali OR “Marshall Islands” OR Mauritania OR Mauritius OR Mexico OR Micronesia OR Moldova OR Mongolia OR Montenegro OR Morocco OR Mozambique OR Myanmar OR Namibia OR Nepal OR Nicaragua OR Niger OR Nigeria OR “North Korea” OR Pakistan OR Palestine OR Panama OR “Papua New Guinea” OR Paraguay OR Peru OR Philippines OR Phillippines OR Philipines OR Phillipines OR Principe OR Rwanda OR Ruanda OR “Saint Lucia” OR “Saint Vincent” OR Samoa OR “Sao Tome” OR Senegal OR Serbia OR “Sierra Leone” OR “Solomon Islands” OR Somalia OR “South Africa” OR “South Sudan” OR “Sri Lanka” OR “St Lucia” OR “St Vincent” OR Sudan OR Surinam OR Suriname OR Swaziland OR Syria OR “Syrian Arab Republic” OR Tajikistan OR Tadzhikistan OR Tadjikistan OR Tadzhik OR Tanzania OR Thailand OR Timor-Leste OR Togo OR Tonga OR Tunisia OR Turkey OR Türkiye OR Turkmen OR Turkmenistan OR Tuvalu OR Uganda OR Ukraine OR Uzbek OR Uzbekistan OR Vanuatu OR Venezuela OR Vietnam OR “Viet Nam” OR “West Bank” OR Yemen OR Zambia OR Zimbabwe) OR TI (“Deprived Countries” OR “Deprived Population” OR “Deprived Populations” OR “Developing Countries” OR “Developing Country” OR “Developing Economies” OR “Developing Economy” OR “Developing Nation” OR “Developing Nations” OR “Developing Population” OR “Developing Populations” OR “Developing World” OR “Global South” OR “LAMI Countries” OR “LAMI Country” OR “Less Developed Countries” OR “Less Developed Country” OR “Less Developed Economies” OR “Less Developed Nation” OR “Less Developed Nations” OR “Less Developed World” OR “Lesser Developed Countries” OR “Lesser Developed Nations” OR LMIC OR LMICS OR “Low GDP” OR “Low GNP” OR “Low Gross Domestic” OR “Low Gross National” OR “Low Income Countries” OR “Low Income Country” OR “Low Income Economies” OR “Low Income Economy” OR “Low Income Nations” OR “Low Income Population” OR “Low Income Populations” OR “Lower GDP” OR “lower gross domestic” OR “Lower Income Countries” OR “Lower Income Country” OR “Lower Income Nations” OR “Lower Income Population” OR “Lower Income Populations” OR “Middle Income Countries” OR “Middle Income Country” OR “Middle Income Economies” OR “Middle Income Nation” OR “Middle Income Nations” OR “Middle Income Population” OR “Middle Income Populations” OR “Poor Countries” OR “Poor Country” OR “Poor Economies” OR “Poor Economy” OR “Poor Nation” OR “Poor Nations” OR “Poor Population” OR “Poor Populations” OR “poor world” OR “Poorer Countries” OR “Poorer Economies” OR “Poorer Economy” OR “Poorer Nations” OR “Poorer Population” OR “Poorer Populations” OR “Third World” OR “Transitional Countries” OR “Transitional Country” OR “Transitional Economies” OR “Transitional Economy” OR “Under Developed Countries” OR “Under Developed Country” OR “under developed nations” OR “Under Developed World” OR “Under Served Population” OR “Under Served Populations” OR “Underdeveloped Countries” OR “Underdeveloped Country” OR “underdeveloped economies” OR “underdeveloped nations” OR “underdeveloped population” OR “Underdeveloped World” OR “Underserved Countries” OR “Underserved Nations” OR “Underserved Population” OR “Underserved Populations” OR Afghanistan OR Albania OR Algeria OR Angola OR Argentina OR “Argentine Republic” OR Armenia OR Azerbaijan OR Bangladesh OR Belarus OR Byelarus OR Belorussia OR Belize OR Benin OR Bhutan OR Bolivia OR Bosnia OR Botswana OR Brazil OR Burma OR “Burkina Faso” OR Burundi OR “Cabo Verde” OR “Cape Verde” OR Cambodia OR Cameroon OR “Central African Republic” OR Chad OR China OR Colombia OR Comoros OR Comores OR Comoro OR Congo OR “Côte dIvoire” OR Cuba OR “Democratic Peoples Republic of Korea” OR Djibouti OR Dominica OR “Dominican Republic” OR “East Timor” OR Ecuador OR Equador OR Egypt OR “El Salvador” OR Eritrea OR Eswatini OR Ethiopia OR Fiji OR Gabon OR Gambia OR Gaza OR Georgia OR “Georgia Republic” OR Georgian OR Ghana OR Grenada OR Grenadines OR Guatemala OR Guinea OR “Guinea Bissau” OR Guinea-Bissau OR Guyana OR Haiti OR Herzegovina OR Hercegovina OR Honduras OR India OR Indonesia OR Iran OR Iraq OR Jamaica OR Jordan OR Kazakhstan OR Kenya OR Kiribati OR Kosovo OR Kyrgyz OR Kirghizia OR Kirghiz OR Kirgizstan OR Kirghizstan OR Kyrgyzstan OR “Lao PDR” OR Laos OR Lebanon OR Lesotho OR Liberia OR Libya OR Macedonia OR Madagascar OR Malawi OR Malay OR Malaya OR Malaysia OR Maldives OR Mali OR “Marshall Islands” OR Mauritania OR Mauritius OR Mexico OR Micronesia OR Moldova OR Mongolia OR Montenegro OR Morocco OR Mozambique OR Myanmar OR Namibia OR Nepal OR Nicaragua OR Niger OR Nigeria OR “North Korea” OR Pakistan OR Palestine OR Panama OR “Papua New Guinea” OR Paraguay OR Peru OR Philippines OR Phillippines OR Philipines OR Phillipines OR Principe OR Rwanda OR Ruanda OR “Saint Lucia” OR “Saint Vincent” OR Samoa OR “Sao Tome” OR Senegal OR Serbia OR “Sierra Leone” OR “Solomon Islands” OR Somalia OR “South Africa” OR “South Sudan” OR “Sri Lanka” OR “St Lucia” OR “St Vincent” OR Sudan OR Surinam OR Suriname OR Swaziland OR Syria OR “Syrian Arab Republic” OR Tajikistan OR Tadzhikistan OR Tadjikistan OR Tadzhik OR Tanzania OR Thailand OR Timor-Leste OR Togo OR Tonga OR Tunisia OR Turkey OR Türkiye OR Turkmen OR Turkmenistan OR Tuvalu OR Uganda OR Ukraine OR Uzbek OR Uzbekistan OR Vanuatu OR Venezuela OR Vietnam OR “Viet Nam” OR “West Bank” OR Yemen OR Zambia OR Zimbabwe))

AND

[2000-2025]/py

**SCOPUS**

(INDEXTERMS({Food security} OR {Food supply}) OR TITLE-ABS({food supply} OR {food desert} OR {food deserts} OR {food insecurity} OR food W/2 access OR food W/2 barriers OR {food availability} OR {food insufficiency} OR food W/2 security OR hunger OR {nutritional insecurity} OR malnutrition OR undernourishment OR {food deprivation} OR {dietary inadequacy} OR {caloric insufficiency} OR {food hardship} OR {food poverty} OR {food swamps} OR {food justice} OR {food access disparities} OR starvation OR {food apartheid} OR undernourished OR {food sovereignty} OR {food affordability} OR {food utilization} OR {food agency} OR {food sustainability}))

AND

(INDEXTERMS(Obesity OR Overweight) OR TITLE-ABS(obesity OR overweight OR {excess weight} OR {excess body weight} OR {body weight} OR {weight gain} OR {high BMI} OR {increased BMI} OR adiposity OR {body mass index} OR {weight status} OR {weight-related health} OR obese))

AND

(TITLE-ABS-KEY({Deprived Countries} OR {Deprived Population} OR {Deprived Populations} OR {Developing Countries} OR {Developing Country} OR {Developing Economies} OR {Developing Economy} OR {Developing Nation} OR {Developing Nations} OR {Developing Population} OR {Developing Populations} OR {Developing World} OR {Global South} OR {LAMI Countries} OR {LAMI Country} OR {Less Developed Countries} OR {Less Developed Country} OR {Less Developed Economies} OR {Less Developed Nation} OR {Less Developed Nations} OR {Less Developed World} OR {Lesser Developed Countries} OR {Lesser Developed Nations} OR LMIC OR LMICS OR {Low GDP} OR {Low GNP} OR {Low Gross Domestic} OR {Low Gross National} OR {Low Income Countries} OR {Low Income Country} OR {Low Income Economies} OR {Low Income Economy} OR {Low Income Nations} OR {Low Income Population} OR {Low Income Populations} OR {Lower GDP} OR {lower gross domestic} OR {Lower Income Countries} OR {Lower Income Country} OR {Lower Income Nations} OR {Lower Income Population} OR {Lower Income Populations} OR {Middle Income Countries} OR {Middle Income Country} OR {Middle Income Economies} OR {Middle Income Nation} OR {Middle Income Nations} OR {Middle Income Population} OR {Middle Income Populations} OR {Poor Countries} OR {Poor Country} OR {Poor Economies} OR {Poor Economy} OR {Poor Nation} OR {Poor Nations} OR {Poor Population} OR {Poor Populations} OR {poor world} OR {Poorer Countries} OR {Poorer Economies} OR {Poorer Economy} OR {Poorer Nations} OR {Poorer Population} OR {Poorer Populations} OR {Third World} OR {Transitional Countries} OR {Transitional Country} OR {Transitional Economies} OR {Transitional Economy} OR {Under Developed Countries} OR {Under Developed Country} OR {under developed nations} OR {Under Developed World} OR {Under Served Population} OR {Under Served Populations} OR {Underdeveloped Countries} OR {Underdeveloped Country} OR {underdeveloped economies} OR {underdeveloped nations} OR {underdeveloped population} OR {Underdeveloped World} OR {Underserved Countries} OR {Underserved Nations} OR {Underserved Population} OR {Underserved Populations} OR Afghanistan OR Albania OR Algeria OR Angola OR Argentina OR {Argentine Republic} OR Armenia OR Azerbaijan OR Bangladesh OR Belarus OR Byelarus OR Belorussia OR Belize OR Benin OR Bhutan OR Bolivia OR Bosnia OR Botswana OR Brazil OR Burma OR {Burkina Faso} OR Burundi OR {Cabo Verde} OR {Cape Verde} OR Cambodia OR Cameroon OR {Central African Republic} OR Chad OR China OR Colombia OR Comoros OR Comores OR Comoro OR Congo OR {Côte d’Ivoire} OR Cuba OR {Democratic People’s Republic of Korea} OR Djibouti OR Dominica OR {Dominican Republic} OR {East Timor} OR Ecuador OR Equador OR Egypt OR {El Salvador} OR Eritrea OR Eswatini OR Ethiopia OR Fiji OR Gabon OR Gambia OR Gaza OR Georgia OR {Georgia Republic} OR Georgian OR Ghana OR Grenada OR Grenadines OR Guatemala OR Guinea OR {Guinea Bissau} OR Guinea-Bissau OR Guyana OR Haiti OR Herzegovina OR Hercegovina OR Honduras OR India OR Indonesia OR Iran OR Iraq OR Jamaica OR Jordan OR Kazakhstan OR Kenya OR Kiribati OR Kosovo OR Kyrgyz OR Kirghizia OR Kirghiz OR Kirgizstan OR Kirghizstan OR Kyrgyzstan OR {Lao PDR} OR Laos OR Lebanon OR Lesotho OR Liberia OR Libya OR Macedonia OR Madagascar OR Malawi OR Malay OR Malaya OR Malaysia OR Maldives OR Mali OR {Marshall Islands} OR Mauritania OR Mauritius OR Mexico OR Micronesia OR Moldova OR Mongolia OR Montenegro OR Morocco OR Mozambique OR Myanmar OR Namibia OR Nepal OR Nicaragua OR Niger OR Nigeria OR “North Korea” OR Pakistan OR Palestine OR Panama OR {Papua New Guinea} OR Paraguay OR Peru OR Philippines OR Phillippines OR Philipines OR Phillipines OR Principe OR Rwanda OR Ruanda OR {Saint Lucia} OR {Saint Vincent} OR Samoa OR {Sao Tome} OR Senegal OR Serbia OR {Sierra Leone} OR {Solomon Islands} OR Somalia OR {South Africa} OR {South Sudan} OR {Sri Lanka} OR {St Lucia} OR {St Vincent} OR Sudan OR Surinam OR Suriname OR Swaziland OR Syria OR {Syrian Arab Republic} OR Tajikistan OR Tadzhikistan OR Tadjikistan OR Tadzhik OR Tanzania OR Thailand OR Timor-Leste OR Togo OR Tonga OR Tunisia OR Turkey OR Türkiye OR Turkmen OR Turkmenistan OR Tuvalu OR Uganda OR Ukraine OR Uzbek OR Uzbekistan OR Vanuatu OR Venezuela OR Vietnam OR {Viet Nam} OR {West Bank} OR Yemen OR Zambia OR Zimbabwe))

AND

PUBYEAR > 1999

**LILACS**

(mh:("food insecurity" OR "food supply") OR ("food supply" OR "food desert" OR "food deserts" OR "food insecurity" OR "food access" OR "food barriers" OR "food availability" OR "food insufficiency" OR "food security" OR hunger OR "nutritional insecurity" OR malnutrition OR undernourishment OR "food deprivation" OR "dietary inadequacy" OR "caloric insufficiency" OR "food hardship" OR "food poverty" OR "food swamps" OR "food justice" OR "food access disparities" OR starvation OR “food apartheid” OR undernourished OR “food sovereignty” OR “food affordability” OR “food utilization” OR “food agency” OR “food sustainability”))

AND

(mh:(obesity OR overweight) OR (obesity OR overweight OR "excess weight" OR "excess body weight" OR "body weight" OR "weight gain" OR "high BMI" OR "increased BMI" OR adiposity OR "body mass index" OR "weight status" OR "weight-related health" OR obese))

AND

(“Deprived Countries” OR “Deprived Population” OR “Deprived Populations” OR “Developing Countries” OR “Developing Country” OR “Developing Economies” OR “Developing Economy” OR “Developing Nation” OR “Developing Nations” OR “Developing Population” OR “Developing Populations” OR “Developing World” OR “Global South” OR “LAMI Countries” OR “LAMI Country” OR “Less Developed Countries” OR “Less Developed Country” OR “Less Developed Economies” OR “Less Developed Nation” OR “Less Developed Nations” OR “Less Developed World” OR “Lesser Developed Countries” OR “Lesser Developed Nations” OR LMIC OR LMICS OR “Low GDP” OR “Low GNP” OR “Low Gross Domestic” OR “Low Gross National” OR “Low Income Countries” OR “Low Income Country” OR “Low Income Economies” OR “Low Income Economy” OR “Low Income Nations” OR “Low Income Population” OR “Low Income Populations” OR “Lower GDP” OR “lower gross domestic” OR “Lower Income Countries” OR “Lower Income Country” OR “Lower Income Nations” OR “Lower Income Population” OR “Lower Income Populations” OR “Middle Income Countries” OR “Middle Income Country” OR “Middle Income Economies” OR “Middle Income Nation” OR “Middle Income Nations” OR “Middle Income Population” OR “Middle Income Populations” OR “Poor Countries” OR “Poor Country” OR “Poor Economies” OR “Poor Economy” OR “Poor Nation” OR “Poor Nations” OR “Poor Population” OR “Poor Populations” OR “poor world” OR “Poorer Countries” OR “Poorer Economies” OR “Poorer Economy” OR “Poorer Nations” OR “Poorer Population” OR “Poorer Populations” OR “Third World” OR “Transitional Countries” OR “Transitional Country” OR “Transitional Economies” OR “Transitional Economy” OR “Under Developed Countries” OR “Under Developed Country” OR “under developed nations” OR “Under Developed World” OR “Under Served Population” OR “Under Served Populations” OR “Underdeveloped Countries” OR “Underdeveloped Country” OR “underdeveloped economies” OR “underdeveloped nations” OR “underdeveloped population” OR “Underdeveloped World” OR “Underserved Countries” OR “Underserved Nations” OR “Underserved Population” OR “Underserved Populations” OR Afghanistan OR Albania OR Algeria OR Angola OR Argentina OR “Argentine Republic” OR Armenia OR Azerbaijan OR Bangladesh OR Belarus OR Byelarus OR Belorussia OR Belize OR Benin OR Bhutan OR Bolivia OR Bosnia OR Botswana OR Brazil OR Burma OR “Burkina Faso” OR Burundi OR “Cabo Verde” OR “Cape Verde” OR Cambodia OR Cameroon OR “Central African Republic” OR Chad OR China OR Colombia OR Comoros OR Comores OR Comoro OR Congo OR “Côte d’Ivoire” OR Cuba OR “Democratic People’s Republic of Korea” OR Djibouti OR Dominica OR “Dominican Republic” OR “East Timor” OR Ecuador OR Equador OR Egypt OR “El Salvador” OR Eritrea OR Eswatini OR Ethiopia OR Fiji OR Gabon OR Gambia OR Gaza OR Georgia OR “Georgia Republic” OR Georgian OR Ghana OR Grenada OR Grenadines OR Guatemala OR Guinea OR “Guinea Bissau” OR Guinea-Bissau OR Guyana OR Haiti OR Herzegovina OR Hercegovina OR Honduras OR India OR Indonesia OR Iran OR Iraq OR Jamaica OR Jordan OR Kazakhstan OR Kenya OR Kiribati OR Kosovo OR Kyrgyz OR Kirghizia OR Kirghiz OR Kirgizstan OR Kirghizstan OR Kyrgyzstan OR “Lao PDR” OR Laos OR Lebanon OR Lesotho OR Liberia OR Libya OR Macedonia OR Madagascar OR Malawi OR Malay OR Malaya OR Malaysia OR Maldives OR Mali OR “Marshall Islands” OR Mauritania OR Mauritius OR Mexico OR Micronesia OR Moldova OR Mongolia OR Montenegro OR Morocco OR Mozambique OR Myanmar OR Namibia OR Nepal OR Nicaragua OR Niger OR Nigeria OR “North Korea” OR Pakistan OR Palestine OR Panama OR “Papua New Guinea” OR Paraguay OR Peru OR Philippines OR Phillippines OR Philipines OR Phillipines OR Principe OR Rwanda OR Ruanda OR “Saint Lucia” OR “Saint Vincent” OR Samoa OR “Sao Tome” OR Senegal OR Serbia OR “Sierra Leone” OR “Solomon Islands” OR Somalia OR “South Africa” OR “South Sudan” OR “Sri Lanka” OR “St Lucia” OR “St Vincent” OR Sudan OR Surinam OR Suriname OR Swaziland OR Syria OR “Syrian Arab Republic” OR Tajikistan OR Tadzhikistan OR Tadjikistan OR Tadzhik OR Tanzania OR Thailand OR Timor-Leste OR Togo OR Tonga OR Tunisia OR Turkey OR Türkiye OR Turkmen OR Turkmenistan OR Tuvalu OR Uganda OR Ukraine OR Uzbek OR Uzbekistan OR Vanuatu OR Venezuela OR Vietnam OR “Viet Nam” OR “West Bank” OR Yemen OR Zambia OR Zimbabwe)

**S3. Search strategy (Spanish)**

**LILACS**

(mh:("inseguridad alimentaria" OR " suministro de alimentos") OR (" suministro de alimentos" OR "desierto alimentario" OR "desiertos alimentarios" OR "inseguridad alimentaria" OR "acceso a los alimentos" OR "barreras alimentarias" OR "disponibilidad de alimentos" OR "insuficiencia alimentaria" OR "seguridad alimentaria" OR hambre OR "inseguridad nutricional" OR desnutrición OR subalimentación OR "privación alimentaria" OR "inadecuación dietética" OR "insuficiencia calórica" OR "dificultades alimentarias" OR "pobreza alimentaria" OR "pantanos alimentarios" OR "justicia alimentaria" OR "disparidades en el acceso a alimentos" OR “soberanía alimentaria” OR “asequibilidad de los alimentos” OR “utilización de los alimentos” OR “agencia alimentaria” OR “sostenibilidad de los alimentos” OR “sostenibilidad alimentaria”))

AND

(mh:(obesidad OR sobrepeso) OR (obesidad OR sobrepeso OR "exceso de peso" OR "exceso de peso corporal" OR "peso corporal" OR "aumento de peso" OR "IMC alto" OR "IMC incrementado" OR adiposidad OR "índice de masa corporal" OR "estado de peso" OR "salud relacionada con el peso" OR obeso OR obesa))

AND

(“Países desfavorecidos” OR “Población desfavorecida” OR “Poblaciónes desfavorecidas” OR “Países en desarrollo” OR “Developing Country” OR “Economías en desarrollo” OR “Developing Economy” OR “Nación en desarrollo” OR “Naciónes en desarrollo” OR “Población en desarrollo” OR “Poblaciónes en desarrollo” OR “Mundo en desarrollo” OR “Países de ingesos bajos y medianos” OR “País de ingeso bajo y mediano” OR “Países menos desarrollados” OR “País menos desarrollado” OR “Economías menos desarrolladas” OR “Nación menos desarrollado” OR “Naciónes menos desarrollados” OR “Mundo menos desarrollado” OR PIBM OR PBMI OR “PIB bajo” OR “PNB bajo” OR “Producto interno bruto bajo” OR “Producto nacional bruto bajo” OR “Países de bajos ingresos” OR “País de bajos ingresos” OR “Economías de bajos ingresos” OR “Economía de bajos ingresos” OR “Naciones de bajos ingresos” OR “Población de bajos ingresos” OR “Poblaciónes de bajos ingresos” OR “PIB más bajo” OR “producto interno bruto más bajo” OR “Países de ingresos más bajos” OR “País de ingresos más bajos” OR “Naciones de ingresos más bajos” OR “Poblacion de ingresos más bajos” OR “Poblaciones de ingresos más bajos” OR “Países de ingresos medianos” OR “País de ingresos medianos” OR “Economías de ingresos medianos” OR “Nación de ingresos medianos” OR “Naciones de ingresos medianos” OR “Población de ingresos medianos” OR “Poblaciones de ingresos medianos” OR “Países pobres” OR “País pobre” OR “Economías pobres” OR “Economía pobre” OR “Nación pobre” OR “Naciones pobres” OR “Población pobre” OR “Poblaciones pobres” OR “mundo pobre” OR “Países más pobres” OR “Economías más pobres” OR “Economía más pobre” OR “Naciones más pobres” OR “Población más pobre” OR “Poblaciones más pobres” OR “Tercer mundo” OR “Países en transición” OR “País en transición” OR “Economías en transición” OR “Economía en transición” OR “Países subdesarrollados” OR “País subdesarrollado” OR “naciones subdesarrolladas” OR “Mundo subdesarrollado” OR “Población desatendida” OR “Poblaciones desatendidas” OR Afghanistan OR Afganistán OR Albania OR Algeria OR Argelia OR Angola OR Argentina OR “República Argentina” OR Armenia OR Azerbaijan OR Azerbaiyán OR Bangladesh OR Bangladés OR Belarus OR Byelarus OR Belorussia OR Bielorrusia OR Belize OR Belice OR Benin OR Bhutan OR Bolivia OR Bosnia OR Botswana OR Botsuana OR Brasil OR Brazil OR Burma OR “Burkina Faso” OR Burundi OR “Cabo Verde” OR “Cape Verde” OR Cambodia OR Camboya OR Cameroon OR Camerún OR “República Centroafricana” OR Chad OR China OR Colombia OR Comoros OR Comores OR Comoro OR Congo OR “Costa de Marfil” OR “Corea del Norte” OR Cuba OR Djibouti OR Dominica OR “Timor del Este” OR Ecuador OR Equador OR Egipto OR “El Salvador” OR Eritrea OR Eswatini OR Etiopía OR Fiji OR Filipinas OR Gabón OR Gambia OR Gaza OR Georgia OR “República de Georgia” OR Georgian OR Ghana OR Granada OR Grenadines OR Guatemala OR Guinea OR “Guinea Bissau” OR Guinea-Bissau OR Guyana OR Haiti OR Herzegovina OR Hercegovina OR Honduras OR India OR Indonesia OR Irán OR Irak OR Jamaica OR Jordania OR Kazajistán OR Kenya OR Kiribati OR Kosovo OR Kyrgyz OR Kirghizia OR Kirghiz OR Kirgizstán OR Kirghizstan OR Kyrgyzstan OR “Lao PDR” OR Laos OR Líbano OR Lesoto OR Liberia OR Libia OR Macedonia OR Madagascar OR Malawi OR Malay OR Malaya OR Malasia OR Maldivas OR Mali OR “Marshall Islands” OR Mauritania OR Mauricio OR México OR Micronesia OR Moldovia OR Mongolia OR Montenegro OR Marruecos OR Mozambique OR Myanmar OR Namibia OR Nepal OR Nicaragua OR Niger OR Nigeria OR Pakistán OR Palestina OR Panamá OR “Papua Nueva Guinea” OR Paraguay OR Perú OR Príncipe OR “República Dominicana” OR Ruanda OR “Saint Lucia” OR “Saint Vincent” OR Samoa OR “Sao Tomé” OR Senegal OR Serbia OR “Sierra Leona” OR “Solomon Islands” OR Somalia OR “Sudáfrica OR “Sudán del Sur” OR “Sri Lanka” OR “St Lucia” OR “St Vincent” OR Sudán OR Surinam OR Suriname OR Suazilandia OR Siria OR “Syrian Arab Republic” OR Tajikistán OR Tadzhikistan OR Tadjikistan OR Tadzhik OR Tanzania OR Tailandia OR Timor-Leste OR Togo OR Tonga OR Tunisia OR Turquía OR Türkiye OR Turkmen OR Turkmenistán OR Tuvalu OR Uganda OR Ucraina OR Uzbek OR Uzbekistán OR Vanuatu OR Venezuela OR Vietnam OR “Viet Nam” OR “West Bank” OR Yemen OR Zambia OR Zimbabue)

**S4. Google Scholar Search Results**

Date Search:

Results from first ten pages, sorted by relevancy (500 results per language; 1000 total)

| **Keywords Searched** | **Number of Results** | **Number of Results Screened** | **Number of Possibly Relevant Results** | **Total Possibly Relevant Results** |
| --- | --- | --- | --- | --- |
| **English** | | | | |
| “Food insecurity” AND (obesity OR overweight) AND (“low-income country* OR “middle-income countr*” OR LMIC) |  |  |  |  |
| “Food insecurity” AND (BMI OR “body mass index”) AND (“developing econom*” OR “low resource”) |  |  |  |  |
| “Nutrition insecurity” OR “dietary inadequacy” AND (obesity OR “weight gain”) AND (Africa OR Asia OR “Latin America”) |  |  |  |  |
| “Food access” OR “food availability” AND (overweight OR obesity) AND “developing countr*” |  |  |  |  |
| (“Food insecurity” OR “poverty”) AND (obesity OR overweight OR “nutrition transition”) AND “emerging econom*” |  |  |  |  |
| **Spanish** | | | | |
| "inseguridad alimentaria" AND (obesidad OR sobrepeso) AND ("país de ingresos bajos" OR "país de ingresos medios" OR PIBM) |  |  |  |  |
| "inseguridad alimentaria" AND (IMC OR "índice de masa corporal") AND ("en desarrollo" OR "bajos recursos") |  |  |  |  |
| ("inseguridad nutricional" OR "inadecuación dietética") AND (obesidad OR "aumento de peso") AND (África OR Asia OR "América Latina" OR Latinoamérica) |  |  |  |  |
| ("acceso a los alimentos" OR "disponibilidad de alimentos") AND (sobrepeso OR obesidad) AND "país en desarrollo" |  |  |  |  |
| ("inseguridad alimentaria" OR pobreza) AND (obesidad OR sobrepeso OR "transición nutricional") AND ("economía emergente" OR “país en desarrollo") |  |  |  |  |

**S5. ProQuest Dissertations & Theses Search Results**

Date Searched:

Results from the first five pages, sorted by relevancy (375 results per language; 750 total)

| **Keywords Searched** | **Number of Results** | **Number of Results Screened** | **Number of Possibly Relevant Results** | **Total Possibly Relevant Results** |
| --- | --- | --- | --- | --- |
| **English** | | | | |
| “Food insecurity” AND (obesity OR overweight) AND (“low-income country* OR “middle-income countr*” OR LMIC) |  |  |  |  |
| “Food insecurity” AND (BMI OR “body mass index”) AND (“developing econom*” OR “low resource”) |  |  |  |  |
| “Nutrition insecurity” OR “dietary inadequacy” AND (obesity OR “weight gain”) AND (Africa OR Asia OR “Latin America”) |  |  |  |  |
| “Food access” OR “food availability” AND (overweight OR obesity) AND “developing countr*” |  |  |  |  |
| (“Food insecurity” OR “poverty”) AND (obesity OR overweight OR “nutrition transition”) AND “emerging econom*” |  |  |  |  |
| **Spanish** | | | | |
| "inseguridad alimentaria" AND (obesidad OR sobrepeso) AND ("país de ingresos bajos" OR "país de ingresos medios" OR PIBM) |  |  |  |  |
| "inseguridad alimentaria" AND (IMC OR "índice de masa corporal") AND ("en desarrollo" OR "bajos recursos") |  |  |  |  |
| ("inseguridad nutricional" OR "inadecuación dietética") AND (obesidad OR "aumento de peso") AND (África OR Asia OR "América Latina" OR Latinoamérica) |  |  |  |  |
| ("acceso a los alimentos" OR "disponibilidad de alimentos") AND (sobrepeso OR obesidad) AND "país en desarrollo" |  |  |  |  |
| ("inseguridad alimentaria" OR pobreza) AND (obesidad OR sobrepeso OR "transición nutricional") AND ("economía emergente" OR “país en desarrollo") |  |  |  |  |
